# Opening the Black Box of Artificial Intelligence for Clinical Decision Support: A Study Predicting Stroke Outcome

**DOI:** 10.1101/19010975

**Authors:** Esra Zihni, Vince Istvan Madai, Michelle Livne, Ivana Galinovic, Ahmed A. Khalil, Jochen B. Fiebach, Dietmar Frey

**Author notes:** These authors contributed equally to this work (Esra Zihni, Vince Istvan Madai).

## Abstract

**Background:** State-of-the-art machine learning (ML) artificial intelligence methods are increasingly leveraged in clinical predictive modeling to provide clinical decision support systems to physicians. Modern ML approaches such as artificial neural networks (ANNs) and tree boosting often perform better than more traditional methods like logistic regression. On the other hand, these modern methods yield a limited understanding of the resulting predictions. However, in the medical domain, understanding of applied models is essential, in particular, when informing clinical decision support. Thus, in recent years, interpretability methods for modern ML methods have emerged to potentially allow explainable predictions paired with high performance.

**Methods:** To our knowledge, we present in this work the first explainability comparison of two modern ML methods, tree boosting and multilayer perceptrons (MLPs), to traditional logistic regression methods using a stroke outcome prediction paradigm. Here, we used clinical features to predict a dichotomized 90 days post-stroke modified Rankin Scale (mRS) score. For interpretability, we evaluated clinical features’ importance with regard to predictions using deep Taylor decomposition for MLP, Shapley values for tree boosting and model coefficients for logistic regression.

**Results:** With regard to performance as measured by AUC values on the test dataset, all models performed comparably: Logistic regression AUCs were 0.82, 0.82, 0.79 for three different regularization schemes; tree boosting AUC was 0.81; MLP AUC was 0.81. Importantly, the interpretability analysis demonstrated consistent results across models by rating age and stroke severity consecutively amongst the most important predictive features. For less important features, some differences were observed between the methods.

**Conclusions:** Our analysis suggests that modern machine learning methods can provide explainability which is compatible with domain knowledge interpretation and traditional method rankings. Future work should focus on replication of these findings in other datasets and further testing of different explainability methods.

## Introduction

Machine learning (ML) techniques are state-of-the-art in predictive modeling in fields like computer vision and autonomous navigation [1]. Increasingly, these tools are leveraged for clinical predictive modeling and clinical decision support, where clinical values are used to predict a clinical status, e.g. a diagnosis, outcome or risk [2,3]. Here, newer machine learning techniques - we will refer to them as modern machine learning techniques in this work - including artificial neural nets (ANN), especially deep learning (DL), and ensemble models such as tree boosting have often shown higher performance than traditional machine learning techniques such as linear or logistic regression, e.g. [4–8].

However, a common criticism of these modern techniques is that while they might increase model performance they do not provide the possibility to explain the resulting predictions [9]. In contrast, traditional techniques allow explanations by various means and this approach has been the backbone of explainable clinical predictive modeling to date [10]. The necessity of interpretable ML systems are of particular concern in the medical domain. An explainable AI system is essential to provide: 1) Interpretation and safe-check of the acquired results during development [11]. 2) Better assessment of safety and fairness of medical products, especially regarding bias, during the regulatory process [12]. 3) Domain knowledge supported interpretation leading to increased trust by the physicians, other healthcare professionals, and patients [12]: Some argue that black box approaches are unacceptable for clinical decision support from the physician’s point-of-view [13] and from the patient’s point-of-view [14]. Thus, currently, researchers and developers are facing an unfortunate trade-off: either to use methods with potentially higher performance or to use methods providing explainability to comply with ethical and regulatory requirements [9].

Fortunately, interpretability methods tailored to modern machine learning algorithms have emerged lately, therefore potentially allowing high performance and explainable models. For one, in the last few years several techniques have been developed to open the most notorious black box, namely artificial neural networks and provide explainable models [11]. Moreover, tree boosting provides high performance clinical predictive modeling and also allow the calculation of feature importance and ranking, e.g. Lundberg et al [15]. However, to our knowledge, these approaches have not yet been compared to the traditional methods in terms of interpretability for clinical predictive modeling.

In the present work, we thus compared the above mentioned two modern ML methods, ANNs and tree boosting, to traditional methods with regard to explainability. We chose a well-characterized stroke clinical outcome paradigm. Here, available clinical features such as age, the severity of the stroke or information about treatment are used to predict the 3 months post-stroke outcome. Many replications in the past have established main factors driving the prediction, namely age and stroke severity, e.g. [16–19]. Thus, within this paradigm, modern machine learning explanations can be interpreted against a baseline. Concretely, we used a multilayer perceptron (MLP) with deep Taylor decomposition as an example for an explainable ANN approach [20], the CATBOOST algorithm with Shapley Additive exPlanations (SHAP) values as an example for explainable tree boosting [15] and compared performance and explainability with different versions of regularized logistic regression for a binary outcome (GLM, LASSO, and Elastic Net).

## Methods

### Patients and clinical metadata pre-processing

In a retrospective analysis, patients with acute ischemic stroke from the 1000plus study were included [18]. The study was approved by the local ethics committee in accordance with the Helsinki declaration and all patients gave written informed consent. Patients were triaged into receiving iv-tissue-plasminogen-activator (tPA) for thrombolysis therapy or conservative therapy. The modified Rankin Scale (mRS), representing the degree of disability or dependence in the daily activities, was assessed for each patient 3 months post-stroke via a telephone call. The available database consisted of 514 patients who received imaging at 3 imaging time points. Of these, 104 were lost-to-follow-up and had no mRS values. 1 patient had to be excluded due to values outside of the possible parameter range. Moreover, 95 patients had to be excluded due to infratentorial stroke and no visible DWI lesions. Specific further inclusion criteria of our sub-study were a ratio of at least 1 to 4 for binary variables (absence/presence) and no more than 5% missing values resulting in the final number of 314 patients and the following clinical parameters for the predictive models: age, sex, initial NIHSS (National Institute of Health Stroke Scale; measuring stroke severity), history of cardiac disease, history of diabetes mellitus, presence of hypercholesterolemia, and thrombolysis treatment. Missing values were imputed using mean imputation. The continuous parameters were centered using zero-mean unit-variance normalization. For a summary of the patients’ clinical features and their distribution, see Table 1.

**Table 1.** Summary of the clinical data. The table summarizes the distribution of the selected clinical data covariates acquired in the acute clinical setting. NIHSS stands for National Institutes of Health Stroke Scale; IQR indicates the interquartile range.

| Clinical information | Value |
| --- | --- |
| Median age (IQR) | 72 (15) |
| Sex (Females/ Males) | 196 / 118 |
| Median initial NIHSS (IQR) | 3 (5) |
| Cardiac history (yes/ no) | 84 / 230 |
| Diabetes mellitus (yes/ no) | 79 / 235 |
| Hypercholesterolemia (yes/ no) | 182 / 132 |
| Thrombolysis (yes / no) | 74 / 240 |

### Data Accessibility

Data cannot be shared publicly because of data protection laws. Data might be available from the institutional ethics committee of Charité Universitätsmedizin Berlin (contact via) for researchers who meet the criteria for access to confidential data. The code used in the manuscript is available on Github (https://github.com/prediction2020/explainable-predictive-models).

### Outcome prediction supervised machine learning framework

In a supervised machine-learning framework, the clinical parameters (Table 1) were used to predict the final outcome of stroke patients in terms of dichotomized 3-months post-stroke mRS, where mRS ϵ {0,1,2} indicates a good outcome (i.e. class label *y*_*i*_ = 0 for a given observation *i*) and mRS ϵ {3,4,5,6} indicates a bad outcome (i.e. class label *y*_*i*_ = 1 for a given observation *i*). The applied dichotomization resulted in 88 positive (i.e. bad outcome) and 226 negative (i.e. good outcome) classes.

### Feature multicollinearity

Importantly, methods for feature ranking can be influenced by feature multicollinearity. Particularly, Beta weights in regression analysis can be erroneous in case of multicollinearity [19,20] and certain applications of feature importance calculation for tree boosting are simplified under the assumption of feature independence. To ensure an unbiased comparison of the models interpretability we estimated multicollinearity of the features using the variance inflation factor (VIF) [21]. The chosen features in the analysis demonstrated negligible multicollinearity with VIFs < 1.91 (Age: 1.15; Sex: 1.91, NIHSS: 1.28; Cardiac history: 1.33; Diabetes: 1.36; Hypercholesterolemia: 1.74; Thrombolysis: 1.50). This makes our stroke outcome paradigm particularly suited to compare explainability.

### Predictive modeling and Interpretability

In this study, machine-learning (ML) methods were applied to predict the final outcome based on clinical data. In the context of tabular data as in the given study, the interpretability of the resulting models corresponds to a rating of feature importance. The interpretability frameworks suggested in this study are tailored to the models and therefore indicate the relative contribution of the features to the respective model prediction. The different ML algorithms and the corresponding interpretability derivations are described as follows.

#### Traditional (linear) ML frameworks

##### 1. Generalized linear model (GLM)

GLM is a generalization of linear regression that allows for a response to be dichotomous instead of continuous. Hence the model predicts the probability of a bad outcome (vs. good outcome) based on a set of explanatory variables according to the following relation:

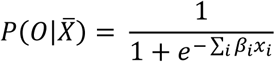

where 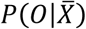 is the probability for a good or a bad outcome (*O* = 0 or *O* = 1 respectively) given the vector of corresponding covariates 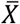. *β* stands for the model parameterization. The objective function for the optimization problem is defined by maximum likelihood estimation (MLE):

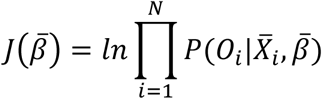

where 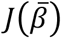 stands for the objective function for the given model parametrization, 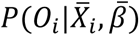 is the predicted outcome probability for the given covariates 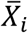 and model parametrization *β* and *N* is the number of observations. In this formulation, this special case of a GLM is also known as logistic regression.

##### 2. Lasso

Lasso, standing for least absolute shrinkage and selection operator, provides the L1 regularized version of GLM. An L1 penalization of the model parametrization reduces overfitting of the model and is applied by the addition of the L1 regularization term to the objective function:

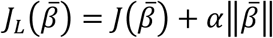

where 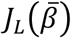 stands for the Lasso objective function and *α* si the scaling factor hyperparameter.

##### 3. Elastic Nets

Similarly to Lasso, elastic net provide a regularized variate of the GLM. Here two types of regularization terms are added to the objective function that provide L1 and L2 penalization of the model parametrization respectively:

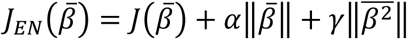

where 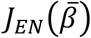 stands for the elastic nets objective function and *α* and *γ* are the scaling factors hyperparameters.

For the three linear models, the interpretability of the models was deduced using the resulted model parametrization. Hence, the rating of the features was derived by the values of the model coefficients *β*. As outlined above, this is sufficient since our features do not exhibit collinearity [20].

#### Modern (nonlinear) ML frameworks

##### 4. Tree boosting (CatBoost)

Treeboosting solves the described classification problem by producing a prediction model as an ensemble of weak classification models, i.e. classifiers. As an ensemble method, the algorithm builds many weak classifiers in the form of decision trees and then integrates them into one cumulative prediction model to obtain better performance than any of the constituent classifiers. The prediction is then given using Kadditive functions:

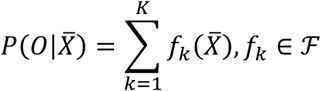

where ℱ = {*f*(*x*) = *w*_*q*(*x*)_}(*q*: ℝ^*m*^ → *T*, w ϵℝ^*T*^) is the space of regression trees. Here *q* denotes the structure of each tree and *T* is the number of leaves in the tree. Each fxrepresents an independent tree structure *q* and leaf weights *w*. The output of the regression trees is a continuous score represented by *w*_*i*_ for leaf *i*. Each observation is classified using each constituent tree to the corresponding leafs and the outcome prediction 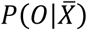 is finally calculated as the cumulative sum of scores of the corresponding leafs. The objective function for optimization constitutes of the convex loss function, here chosen as logistic function, and a regularization component:

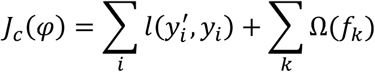

where the convex loss is given by :

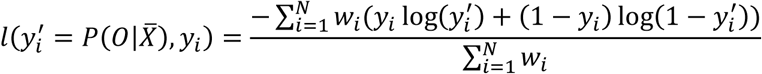

and the regularization component is given by :

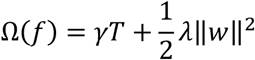

Here *φ* represents the corresponding model parametrization. In this study we used the CATBOOST module to implement the tree boosting model allowing to successfully integrate both numerical and categorical features [22].

In the context of tree boosting models, SHapley Additive exPlanations (SHAP) values construct a robust unified interpretability framework, breaking down the prediction to show the impact of each input feature [23]. The SHAP values attribute to each feature the corresponding change in the model prediction when conditioning on that feature, compared with the prediction with an unknown feature input. The overall rating of the feature contribution to the model is then achieved by aggregating the SHAP values over all observations.

##### 5. MLP

A multilayer perceptron (MLP) is a type of feedforward artificial neural network that is composed of connectionist neurons, also known as perceptrons, in a layered structure. An MLP architecture is constructed of 3 components: 1) an input layer to receive the information 2) an output layer that makes a decision or prediction about the input and 3) one or more hidden layers that allow for feature extraction and modeling of the covariates dynamics using nonlinear transformations. According to the universal approximation theorem, an MLP with one hidden layer (see Figure 1) can approximate any function [24].

**Fig 1.**
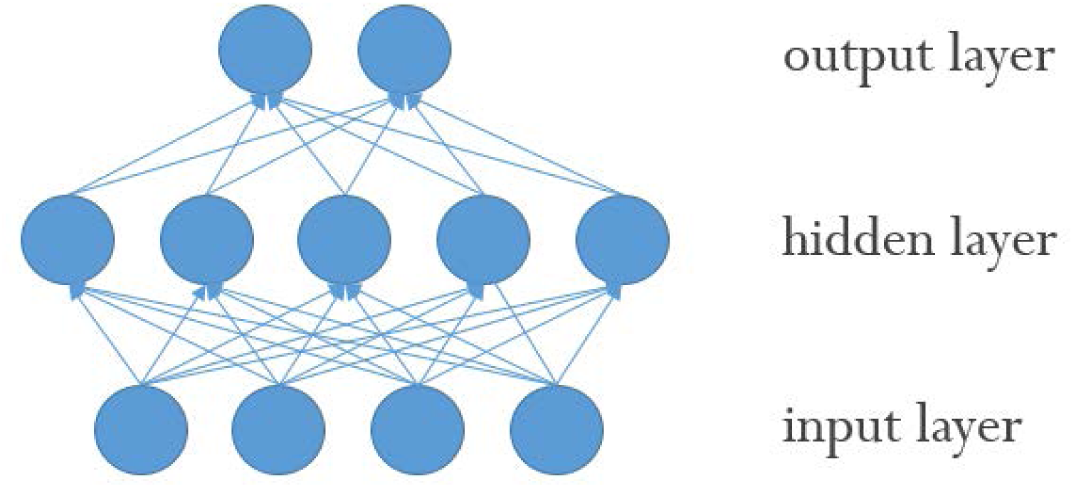
Illustration of an MLP with a single hidden layer. Graphical representation of a fully connected MLP network with a single hidden layer. The arrows represent the direction of the information flow.

Here the model prediction is given by:

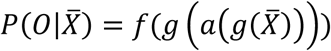

where 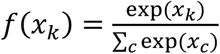 the (softmax) output layer activation, *k* is the predicted class and *c* is any of the possible classes for prediction. *a*(*x*) = max(0, *x*) denotes the hidden layer activation function 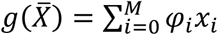 where *M* represents the number of nodes in the layer.

The core objective function utilized for the MLP model was binary cross-entropy:

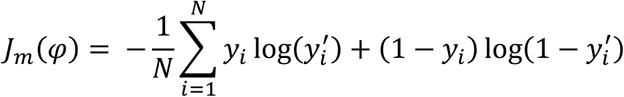

where *φ* represents the corresponding model parametrization. Regularization of the model was entailed using: 1) L1 regularization, i.e. linear penalization of the model parametrization 2) dropout, i.e. random drop of nodes at each stage of the training process with a probabilistic rate *DR* and consecutive weighting of each of the nodes’ output with *(1-DR)* in the prediction inference to yield the expected value of the output.

Several gradient based algorithms were proposed in the recent years as a means to interpret deep neural networks, among them as saliency maps, SmoothGrad, LRP and others, e.g. [25–27]. In the present work, deep Taylor decomposition was chosen as a robust implementation over different data types and neural network architectures [28], which is a recommended technique [10]. The overall features importance was calculated as the weighted average of the observations with relation to the confidence of prediction:

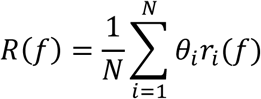

with 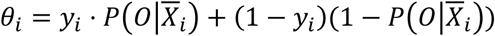 where *R(f)* is the normalized feature rating and *r*_*i*_*(f)* is the feature contribution for the given MLP model for observation *i* using deep Taylor decomposition.

### Models training and validation

The data were randomly split into training- and test sets with a corresponding 4:1 ratio. To target class imbalance the training set was randomly sub-sampled to yield uniform class distribution. The models were then tuned using 10-folds cross-validation. The whole process was repeated 50 times (shuffles). Table 2 provides a summary of the tuned hyperparameters for each model.

**Table 2.** Summary of hyperparameters tuning. The table details the hyperparameters and corresponding range that were tuned for each model in the cross-validation process.

| Model | Hyperparameter | Range |
| --- | --- | --- |
| LASSO | C (inverse of regularizer multiplier) | numpy.linspace (0.1,1000,50) |
| Elastic net | L1 ratio | 0, 0.05, 0.1, 0.15, 0.2, 0.25, 0.3, 0.35, 0.4, 0.45, 0.5, 0.55, 0.6, 0.65, 0.7, 0.75, 0.8, 0.85, 0.9, 0.95 |
|  | Alpha | 0.00001, 0.00004, 0.00016, 0.0006, 0.0025, 0.01, 0.04, 0.16, 0.63, 2.5, 10 |
| CatBoost | Tree depth | 2, 4 |
|  | Learning rate | 0.03, 0.1, 0.3 |
|  | Bagging temperature | 0.6, 0.8, 1. |
|  | L2 leaf regularization | 3, 10, 100, 500 |
|  | Leaf estimation iterations | 1, 2 |
| MLP | Number of hidden neurons | 5, 10, 15, 20 |
|  | Learning rate | 0.001, 0.01 |
|  | Batch size | 16, 32 |
|  | Dropout rate | 0.1, 0.2 |
|  | L1 regularization ratio | 0.0001, 0.001 |

### Performance assessment

The model performance was tested on the test set using receiver-operating-characteristic (ROC)-analysis by measuring the area-under-the-curve (AUC). The performance measure was taken as the median value over the number of shuffles.

### Interpretability assessment

The absolute values of the calculated feature importance scores were normalized, i.e. scaled to unit norm, in order to provide comparable feature rating across models: For each sample (each of the 50 shuffles) the calculated importance scores were rescaled to be confined within the range [0,1] with their sum equal to one. Then, for each feature the mean and standard deviation over the samples (shuffles) were calculated and reported as the final rating measures.

## Results

### Performance Evaluation

All models demonstrated comparable performance for 3 months dichotomized mRS prediction as measured by AUC values on the test set: GLM 0.82, Lasso 0.82, Elastic-nets 0.79, Catboost 0.81 and MLP 0.81. While Catboost showed the highest performance, the difference to the other models was very small. For a graphical representation of the models performance on the training and test sets please see Figure 2.

**Fig 2.**
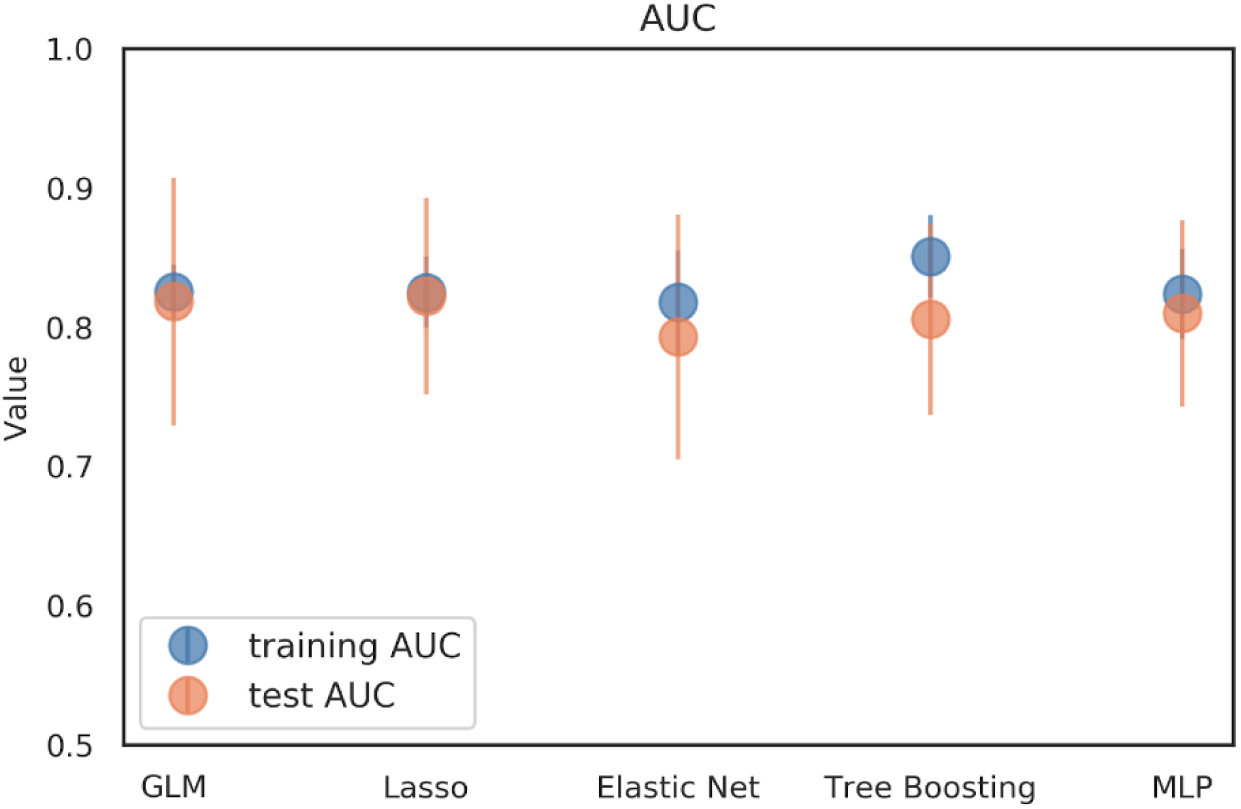
Graphical representation of the model performance results. The graph illustrates the performance of the different models evaluated on the test (blue) and training (orange) sets: generalized linear model (GLM), Lasso, Elastic net, Tree Boosting and multilayer perceptron (MLP). The markers show showing the median AUC over 50 shuffles and the error bars represent interquartile range (IQR). All models showed a similar median AUC around 0.82. The largest difference in performance between training and test set, indicating potential overfitting, was observed for the Catboost model.

### Interpretability analysis

The interpretability analysis demonstrated consistent results across models. All explainable models rated age and initial NIHSS consistently amongst the most important features. The most similar ratings were obtained between the Elastic net and the tree boosting model. The lowest variance amongst feature importance was found for the MLP model. A graphical representation of the results can be found in Figure 3.

**Fig 3.**
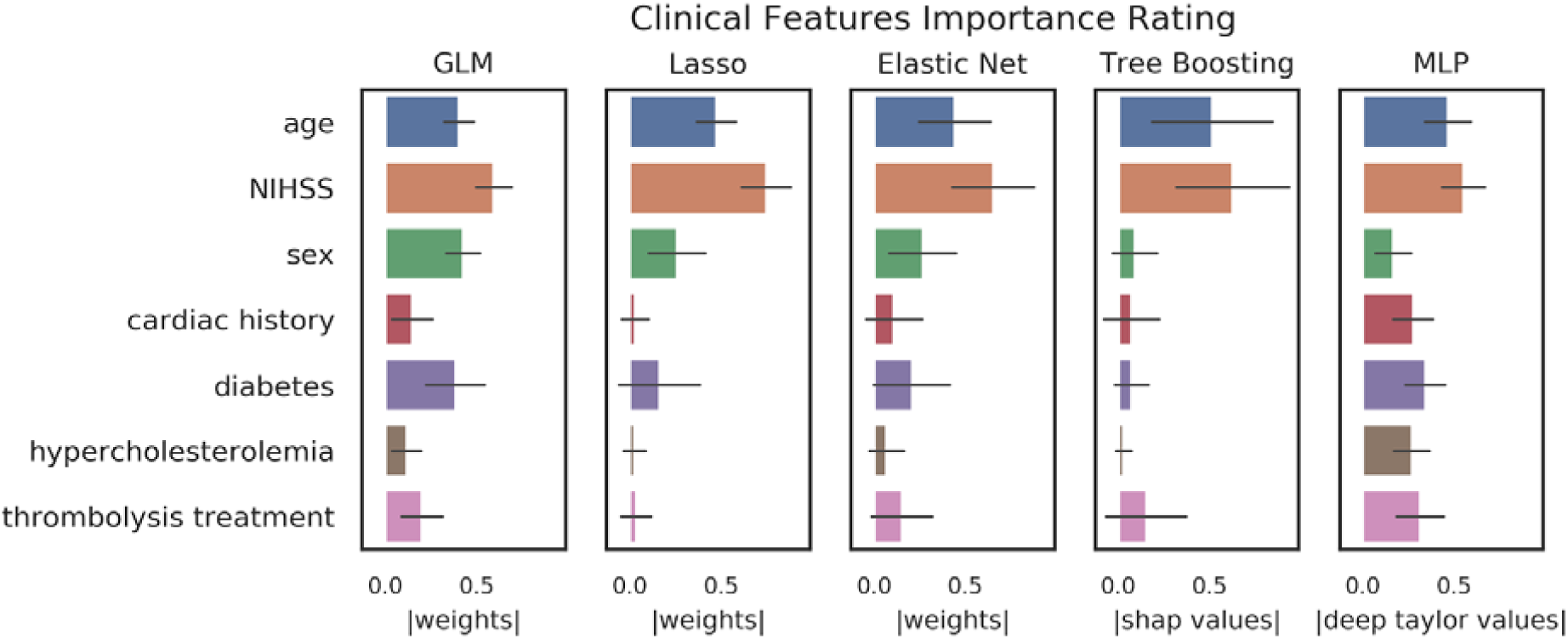
Graphical representation of the feature importance. The figure illustrates the features rating derived from the model-tailored interpretability methods for generalized linear model (GLM), Lasso, Elastic net, Catboost and multilayer perceptron (MLP). For logistic regression techniques the results are given in weights, for Catboost in Shap(ley) values and for MLP in deep Taylor values that were normalized to the range [0,1]. The bar heights represent means and error bars represent standard deviation over samples (shuffles).

## Discussion

In the present work, we have used a well-characterized clinical stroke outcome prediction paradigm to compare the ability of modern and traditional machine learning methods to provide explainability of their predictions. In the context of the presented study, both types of ML methods (artificial neural nets and tree boosting) showed comparable performance and similar interpretability patterns for the most important predictors. We corroborated that modern techniques are not necessarily black boxes, but are able to provide a reliable assessment of feature importance comparable to their traditional counterparts for clinical prediction models. In contrast to other domains, models in healthcare require higher levels of safety given that patients’ life and health is at stake [11]. Here, the explainability of the predictions is a highly important criterion to enable it. Unfortunately, explainability in the modeling context is an ill-defined term than can also have other meanings and several other terms such as interpretability and transparency are in use, e.g. Roscher et al. [29]. We thus stress that standardization of terms is highly needed to facilitate the discussion about explainable AI systems. In the presented work, explainability is mainly examined from a clinical point-of-view, highlighting the ability of humans to understand which clinical features drive the prediction. This is important, as a major goal of clinical predictive modeling is the development of clinical decision support systems (CDSS) aiding healthcare professionals in their clinical decision making, predicting diagnoses, risks, and outcomes [2]. Here, it is important to keep in mind that the requirements for CDSSs go far beyond the model performance [12]. It is established that CDSSs for the clinical setting need to exhibit proven safety [12]. A crucial part of the safety assessment of ML/AI products is to understand why they do what they do, but, more importantly, to understand why and when they might *not* do what is intended. This is important in the light of the increasing awareness of potential biases in models used for healthcare discriminating based on for example sex and gender or ethnicity[30]. Another reason is automation bias - an established cognitive bias - where users tend to believe what a machine is outputting without reflecting on the output [2]. Providing model explainability might mitigate this bias. Thus, it is very likely that future regulatory requirements, e.g. by European MDR and US FDA, will include requests for explainability [31]. Here, our results are highly encouraging. Modern ML methods that are able to provide the potentially highest performance can be combined with methods of explainability and the results are comparable to the established methods for traditional techniques. Thus, researchers and developers are no longer faced with the potential trade-off between lower performance vs. explainability.

However, not only regulatory bodies will require explainability. From the physician point-of-view, black-box approaches might be unacceptable [12,32]. Clinical guidelines for CDSS may therefore profit from explainable predictions. While it has been argued that we have accepted similar uncertainty in medical decision making to date and accuracy alone can be sufficient [33], we would argue that explainability is a must-have when it can be added without limiting the accuracy, as our results suggest. Nonetheless, explainability is a supportive tool and is not a substitute for rigorous clinical validation of any CDSS [33].

We have focused in our work on two promising techniques, namely artificial neural nets and tree boosting. ANNs have shown highly promising results in several areas of healthcare such as medical imaging, information extraction from medical texts and electronic health records, and combining several types of input into one predictive model [5]. Also tree boosting has shown high performance across several medical domains [34]. Tree boosting algorithms are also much easier to train than artificial neural nets and their performance is quite immune to feature scaling and collinearity issues. Another major advantage of tree boosting in healthcare is scalability [35] and thus it is also suited for big data analytics, for example data mining from electronic health records (EHR). Here, tree boosting can achieve comparable performance to deep learning techniques [36]. As evidenced by the above, tree boosting and ANNs represent very versatile and well performing modern ML algorithms in healthcare. Thus, our work is of high practicality for future research and for clinical decision support development.

The main focus of our work was the comparison of explainability in a well-characterized prediction paradigm and not a comparison of performance. It is not surprising that both the traditional and the modern ML methods achieved comparable performance in our dataset. Given the simplicity of the classification problem and the limited dataset, traditional methods are sufficient to capture the relationship of the features to the prediction and complex methods may easily result in overfitting. It is, however, important to note that interpretability without a certain performance level is meaningless: A randomly classifying classifier cannot provide reliable feature importance. Thus, the simplicity of the paradigm we chose is well suited to compare explainability, as the performance is comparable and feature ratings provide a straight-forward result that can be assessed against domain knowledge. Had the performance varied considerably, interpretation of the rankings might have been severely impaired. With regard to our explainability analysis, several more observations are noteworthy. As there is no gold-standard to interpret rankings it can only be performed against domain-knowledge and through replication studies. While we know from previous studies that age, NIHSS and thrombolysis are important predictors to predict stroke outcome (with age and NIHSS being the two strongest) [14–17], it is crucial to include the specifics of the dataset into the interpretation. The median NIHSS of the sample was only 3 and only around 31% of patients received thrombolysis, meaning that many of the patients had smaller - less serious - stroke events. As a consequence, the potential effect of thrombolysis is limited in our sample. Thus we would - like in the above mentioned previous works - expect that age and NIHSS drive the prediction. And indeed, all rankings gave these two very high importance, with the exception of the GLM ranking they were the two most important predictors. The ranking of the lesser predictors, however, varied relatively strongly. Interestingly, elastic net provided the ranking which is most similar to the one provided by tree boosting. From a domain perspective, the most reliable and complete ranking was provided by the tree boosting model, ranking age and NIHSS unequivocally on top, with thrombolysis being slightly more important than the other features. While the MLP gave age and NIHSS the expected high importance, it ranked the presence of diabetes similarly strong. A similar ranking for diabetes can also be observed in the logistic regression models. Although diabetes is known to be an important predictor for bad stroke outcome [37], a feature importance score that is at the same level as NIHSS and age is unexpected. Another striking difference is the high relative importance given to sex by the logistic regression models, which is absent in the rankings provided by the modern methods. Taken together, we observed promising consistent findings, where all methods corroborated the importance of age and NIHSS for stroke outcome prediction. At the same time, we saw distinct differences for diabetes and sex which cannot be explained sufficiently at the current time point. In light of these findings, we certainly do not claim that the explanations provided by the modern methods should be taken without further validation. Our work established that rankings can be obtained for modern machine learning methods and that these rankings are compatible with clinical interpretation, especially regarding the main predictors. The differences between the rankings, however, must be the subject of further research. Here, it must be mentioned that for ANNs multiple other methods than Taylor decomposition exist, which should also be further tested in the future - a task which was beyond the scope of the current work.

Given the aforementioned trade-off between performance and explainability, a distinction between traditional and modern techniques seems justifiable. It carries with it, however, the risk that modern methods are overhyped and used where traditional techniques might perform best. As our results suggest that also modern techniques provide explainability, we would argue that this distinction is irrelevant. Once all important methods for clinical predictive modeling provide validated feature importance we should simply choose the method which seems best suited for the prediction task at hand. We believe that this will greatly facilitate the development of clinical decision support systems.

Our work has several limitations. First, we used only one dataset. Here, our results are promising, but clearly more analyses are warranted to compare rankings provided by modern ML methods with rankings provided by traditional ML methods. Second, to allow comparison with traditional methods, we used a paradigm that utilizes only clinical values. We encourage future works evaluating explainability provided for other data modalities such as imaging.

## Conclusions

For the first time, we established in an empirical analysis on clinical data that modern machine learning methods can provide explainability which is compatible with domain knowledge interpretation and traditional method rankings. This is highly encouraging for the development of explainable clinical predictive models. Future work should validate the explainability methods, further explore the differences between them, and test different predictive modeling frameworks including multiple modalities.

## Data Availability

Data cannot be shared publicly because of data protection laws. Data might be available from the institutional ethics commitee of Charité Universitätsmedizin Berlin (contact via) for researchers who meet the criteria for access to confidential data.

## Declarations

### Disclosures

The authors report no disclosures.

### Funding

This work has received funding by the German Federal Ministry of Education and Research through (1) the grant Centre for Stroke Research Berlin and (2) a Go-Bio grant for the research group PREDICTioN2020 (lead: DF). For open access publication, we acknowledge support from the German Research Foundation (DFG) via the Open Access Publication Fund of Charité - Universitätsmedizin Berlin.

## Notes

### Competing Interest Statement

The authors have declared no competing interest.

## References

1. Khamparia A, Singh KM. A systematic review on deep learning architectures and applications. Expert Syst. 2019;36: e12400. doi:10.1111/exsy.12400

2. Challen R, Denny J, Pitt M, Gompels L, Edwards T, Tsaneva-Atanasova K. Artificial intelligence, bias and clinical safety. BMJ Qual Saf. 2019;28: 231–237. doi:10.1136/bmjqs-2018-008370

3. Ashrafian H, Darzi A. Transforming health policy through machine learning. PLOS Med. 2018;15: e1002692. doi:10.1371/journal.pmed.1002692

4. Miotto R, Wang F, Wang S, Jiang X, Dudley JT. Deep learning for healthcare: review, opportunities and challenges. Brief Bioinform. 2018;19: 1236–1246. doi:10.1093/bib/bbx044

5. Esteva A, Robicquet A, Ramsundar B, Kuleshov V, DePristo M, Chou K, et al. A guide to deep learning in healthcare. Nat Med. 2019;25: 24–29. doi:10.1038/s41591-018-0316-z

6. Luo L, Li J, Liu C, Shen W. Using machine-learning methods to support health-care professionals in making admission decisions. Int J Health Plann Manage. 2019;34: e1236–e1246. doi:10.1002/hpm.2769

7. Jhee JH, Lee S, Park Y, Lee SE, Kim YA, Kang S-W, et al. Prediction model development of late-onset preeclampsia using machine learning-based methods. PLOS ONE. 2019;14: e0221202. doi:10.1371/journal.pone.0221202

8. Livne M, Boldsen JK, Mikkelsen IK, Fiebach JB, Sobesky J, Mouridsen K. Boosted Tree Model Reforms Multimodal Magnetic Resonance Imaging Infarct Prediction in Acute Stroke. Stroke. 2018;49: 912–918. doi:10.1161/STROKEAHA.117.019440

9. Adadi A, Berrada M. Peeking Inside the Black-Box: A Survey on Explainable Artificial Intelligence (XAI). IEEE Access. 2018;6: 52138–52160. doi:10.1109/ACCESS.2018.2870052

10. Nathans LL, Oswald FL, Nimon K. Interpreting Multiple Linear Regression: A Guidebook of Variable Importance. 2012;17: 19.

11. Montavon G, Samek W, Müller K-R. Methods for interpreting and understanding deep neural networks. Digit Signal Process. 2018;73: 1–15. doi:10.1016/j.dsp.2017.10.011

12. Ahmad MA, Eckert C, Teredesai A. Interpretable Machine Learning in Healthcare. Proceedings of the 2018 ACM International Conference on Bioinformatics, Computational Biology, and Health Informatics. New York, NY, USA: ACM; 2018. pp. 559–560. doi:10.1145/3233547.3233667

13. Shortliffe EH, Sepúlveda MJ. Clinical Decision Support in the Era of Artificial Intelligence. JAMA. 2018;320: 2199–2200. doi:10.1001/jama.2018.17163

14. Vayena E, Blasimme A, Cohen IG. Machine learning in medicine: Addressing ethical challenges. PLOS Med. 2018;15: e1002689. doi:10.1371/journal.pmed.1002689

15. Lundberg SM, Lee S-I. A Unified Approach to Interpreting Model Predictions. In: Guyon I, Luxburg UV, Bengio S, Wallach H, Fergus R, Vishwanathan S, et al., editors. Advances in Neural Information Processing Systems 30. Curran Associates, Inc.; 2017. pp. 4765–4774. Available: http://papers.nips.cc/paper/7062-a-unified-approach-to-interpreting-model-predictions.pdf

16. Khosla A, Cao Y, Lin CC-Y, Chiu H-K, Hu J, Lee H. An integrated machine learning approach to stroke prediction. Proceedings of the 16th ACM SIGKDD international conference on Knowledge discovery and data mining. ACM; 2010. pp. 183–192.

17. Asadi H, Dowling R, Yan B, Mitchell P. Machine Learning for Outcome Prediction of Acute Ischemic Stroke Post Intra-Arterial Therapy. PLOS ONE. 2014;9: e88225. doi:10.1371/journal.pone.0088225

18. Weimar C, Roth MP, Zillessen G, Glahn J, Wimmer MLJ, Busse O, et al. Complications following Acute Ischemic Stroke. Eur Neurol. 2002;48: 133–140. doi:10.1159/000065512

19. Parsons MW, Christensen S, McElduff P, Levi CR, Butcher KS, De Silva DA, et al. Pretreatment diffusion-and perfusion-MR lesion volumes have a crucial influence on clinical response to stroke thrombolysis. J Cereb Blood Flow Metab Off J Int Soc Cereb Blood Flow Metab. 2010 [cited 16 Feb 2010]. doi:10.1038/jcbfm.2010.3

20. Montavon G, Lapuschkin S, Binder A, Samek W, Müller K-R. Explaining nonlinear classification decisions with deep Taylor decomposition. Pattern Recognit. 2017;65: 211–222. doi:10.1016/j.patcog.2016.11.008

21. Hotter B, Pittl S, Ebinger M, Oepen G, Jegzentis K, Kudo K, et al. Prospective study on the mismatch concept in acute stroke patients within the first 24 h after symptom onset −1000Plus study. BMC Neurol. 2009;9: 60. doi:10.1186/1471-2377-9-60

22. Nimon KF, Oswald FL. Understanding the Results of Multiple Linear Regression: Beyond Standardized Regression Coefficients. Organ Res Methods. 2013;16: 650–674. doi:10.1177/1094428113493929

23. Miles J. Tolerance and Variance Inflation Factor. Wiley StatsRef: Statistics Reference Online. American Cancer Society; 2014. doi:10.1002/9781118445112.stat06593

24. catboost: Catboost Python Package. Available: https://catboost.ai

25. Csáji BC. Approximation with Artificial Neural Networks. 2001; 45.

26. Smilkov D, Thorat N, Kim B, Viégas F, Wattenberg M. SmoothGrad: removing noise by adding noise. ArXiv170603825 Cs Stat. 2017 [cited 2 Sep 2019]. Available: http://arxiv.org/abs/1706.03825

27. Bach S, Binder A, Montavon G, Klauschen F, Müller K-R, Samek W. On Pixel-Wise Explanations for Non-Linear Classifier Decisions by Layer-Wise Relevance Propagation. PLOS ONE. 2015;10: e0130140. doi:10.1371/journal.pone.0130140

28. Simonyan K, Vedaldi A, Zisserman A. Deep Inside Convolutional Networks: Visualising Image Classification Models and Saliency Maps. ArXiv13126034 Cs. 2013 [cited 2 Sep 2019]. Available: http://arxiv.org/abs/1312.6034

29. Roscher R, Bohn B, Duarte MF, Garcke J. Explainable Machine Learning for Scientific Insights and Discoveries. ArXiv190508883 Cs Stat. 2019 [cited 3 Sep 2019]. Available: http://arxiv.org/abs/1905.08883

30. Nelson GS. Bias in Artificial Intelligence. N C Med J. 2019;80: 220–222. doi:10.18043/ncm.80.4.220

31. johner-institut/ai-guideline. In: GitHub [Internet]. [cited 4 Oct 2019]. Available: https://github.com/johner-institut/ai-guideline

32. Yu K-H, Kohane IS. Framing the challenges of artificial intelligence in medicine. BMJ Qual Saf. 2019;28: 238–241. doi:10.1136/bmjqs-2018-008551

33. London AJ. Artificial Intelligence and Black-Box Medical Decisions: Accuracy versus Explainability. Hastings Cent Rep. 2019;49: 15–21. doi:10.1002/hast.973

34. Zhang Z, Zhao Y, Canes A, Steinberg D, Lyashevska O, AME Big-Data Clinical Trial Collaborative Group W on BO. Predictive analytics with gradient boosting in clinical medicine. Ann Transl Med. 2019;7. doi:10.21037/24543

35. Chen T, Guestrin C. XGBoost: A Scalable Tree Boosting System. ArXiv160302754 Cs. 2016; 785–794. doi:10.1145/2939672.2939785

36. Zhao J, Feng Q, Wu P, Lupu RA, Wilke RA, Wells QS, et al. Learning from Longitudinal Data in Electronic Health Record and Genetic Data to Improve Cardiovascular Event Prediction. Sci Rep. 2019;9: 1–10. doi:10.1038/s41598-018-36745-x

37. Lau L-H, Lew J, Borschmann K, Thijs V, Ekinci EI. Prevalence of diabetes and its effects on stroke outcomes: A meta-analysis and literature review. J Diabetes Investig. 2019;10: 780–792. doi:10.1111/jdi.12932

